# NOTCH3 cysteine variants alter the white matter lesion-brain volume link in the general population

**DOI:** 10.64898/2026.10.04.26364679

**Authors:** Matthieu Pluntz, Jessica Lebenberg, Hugues Chabriat, Anne-Louise Leutenegger

## Abstract

White matter hyperintensities on brain magnetic resonance imaging are established markers of cerebral small vessel disease and typically associate inversely with brain volume in the general population, reflecting irreversible tissue loss. In Cerebral Autosomal Dominant Arteriopathy with Subcortical Infarcts and Leukoencephalopathy, a monogenic small vessel disease caused by NOTCH3 cysteine-altering mutations, white matter hyperintensities may instead reflect tissue water accumulation and perivascular space dilation rather than net structural damage. Whether this distinct pathophysiological relationship is detectable at population level among carriers of NOTCH3 cysteine-altering variants without clinically manifest disease remains unknown.

We conducted a cross-sectional neuroimaging study using UK Biobank data acquired between 2014 and 2023. Among 44,943 participants with complete brain magnetic resonance imaging data (mean age 64 years ± 8 years), 96 carried heterozygous NOTCH3 cysteine-altering variants within epidermal growth factor-like repeat domains 7 to 34 and 44,847 were non-carriers. White matter hyperintensity volume was automatically quantified and normalized to total intracranial volume. Brain parenchymal fraction was defined as combined gray and white matter volumes divided by total intracranial volume. Lacunes were manually assessed in all carriers. Linear regression and generalized additive models examined the association between white matter hyperintensity volume and brain parenchymal fraction stratified by genotype, adjusting for age, sex, lacune count, and vascular risk factors.

NOTCH3 variant carriers showed markedly higher normalized white matter hyperintensity volume than non-carriers (0.83% versus 0.36%; P<10⁻⁸) but only marginally higher brain parenchymal fraction (81.1% versus 80.5%; P=0.03). In non-carriers, white matter hyperintensity volume was inversely associated with brain parenchymal fraction (β=-0.116% per 1% increase; P=3×10⁻⁶). In contrast, NOTCH3 variant carriers demonstrated a positive association (β=+0.51%; P=0.019), strengthened after adjusting for lacune count (β=+0.61%; P=0.006). A significant genotype-specific interaction confirmed this divergence (P=0.036), persisting across age groups and after vascular risk factor adjustment.

NOTCH3 cysteine-altering variant carriers identified at population level exhibit a fundamentally distinct white matter hyperintensity-brain volume relationship compared with non-carriers, consistent with lesions reflecting tissue water accumulation rather than irreversible structural loss at this early, predominantly preclinical disease stage. These findings establish that genetic context modifies neuroimaging biomarker interpretation in cerebral small vessel disease and underscore the need for genotype-aware approaches when evaluating white matter hyperintensities as markers of disease severity or progression.

## Introduction

White matter hyperintensities (WMH) on T2-weighted fluid-attenuated inversion recovery (T2-FLAIR) Magnetic Resonance Imaging (MRI) are a hallmark of cerebral small vessel disease (cSVD) and increase substantially with aging and vascular risk factors.^1^ In population-based studies, WMH burden is consistently associated with cognitive impairment, gait disturbances, and increased risk of stroke and dementia.^2,3^ As a result, WMH volume is widely used as a biomarker of cSVD severity and progression.

At tissue level, WMH could actually reflect heterogeneous pathological substrates, including demyelination, axonal loss, gliosis, or increased interstitial water content.^4,5^ In sporadic cSVD, they are commonly interpreted as markers of chronic ischemic injury leading to irreversible structural brain damage. Consistent with this framework, multiple studies in the general population have demonstrated a robust inverse relationship between WMH volume and global brain volume, supporting the concept that WMH accumulation accelerates age-related brain atrophy.^6^ ^7^ ^8^

This prevailing model was recently challenged by findings obtained in Cerebral Autosomal Dominant Arteriopathy with Subcortical Infarcts and Leukoencephalopathy (CADASIL), the most frequent monogenic form of cSVD caused by heterozygous missense mutations in the NOTCH3 gene. Such mutations lead to an odd number of cysteine residues within one of the 34 epidermal growth factor-like repeat (EGFR) domains of the Notch3 protein,^9^ leading to protein accumulation and vascular dysfunction.^10^ In symptomatic CADASIL patients, Yao et al reported a positive association between WMH volume and brain parenchymal fraction (BPF: total brain volume divided by intracranial volume).^11^ This association emerged only after accounting for lacunes or in their absence,^11^ suggesting that, beyond lacunar-driven tissue loss, WMH burden in CADASIL may not uniformly reflect net structural tissue loss. Neuropathological studies showed that white matter lesions in CADASIL are heterogeneous, combining myelin and axonal loss with dilation of perivascular spaces and extracellular matrix alterations.^12,13^ Imaging studies established that lacunes are the principal contributors to irreversible brain volume loss in CADASIL.^14^ WMH topography might also be critical: periventricular WMH are closely associated with tissue rarefaction and axonal loss, whereas juxtacortical and superficial WMH are likely related to increased interstitial water content and perivascular space dilation.^15,16^ This may also explain why, in CADASIL, even extensive WMH may exert a relatively limited clinical impact in CADASIL, in the absence of overt tissue-destructive lesions.^15^

Recent large population-based genomic studies suggest these observations extend beyond clinically manifest CADASIL, as cysteine-altering NOTCH3 variants are far more common in the general population than previously recognized.^17,18^ In the UK Biobank, carriers of such variants can also exhibit WMH on brain MRI, most often without any overt clinical manifestation.^19^ These findings raise the hypothesis that WMH detected in carriers of NOTCH3 cysteine-altering variants may also share pathophysiological features of symptomatic CADASIL patients with WMH, including a predominance of tissue water accumulation at cerebral level.

To test this hypothesis, we examined the association between WMH volume and BPF in a large population-based cohort from the UK Biobank, stratified by the presence or absence of NOTCH3 cysteine-altering (NOTCH3-Cys) variants. We further assessed, when available, the impact of adjusting for lacunes and explored interactions among age, sex, WMH burden, and NOTCH3 genotype to disentangle the respective contributions of water tissue inflation or of cerebral tissue loss in cSVD.

## Material and Methods

### Study Population and CADASIL Mutation Identification

The UK Biobank (UKB) is a large-scale, population-based prospective cohort comprising 501,941 participants aged 40-69 years at recruitment.^20^ ^21^ All UKB participants provided written informed consent, and the study was approved by the Northwest Multi-Centre Research Ethics Committee. Genetic data were made available to participants only upon explicit request through the UKB’s approved feedback pathway.^22^

Between 2014 and 2023, a subset of 47,486 participants underwent brain MRI. Compared with those without MRI data, scanned participants had a more favorable vascular risk profile (lower hypercholesterolemia, hypertension, diabetes, smoking) but higher daily alcohol intake, and less prior stroke (0.7% vs 1.7%; see Table S1a).

For this study, we analyzed two specific populations with cerebral MRI data: (1) individuals carrying a cysteine NOTCH3 mutation previously reported in CADASIL, and (2) individuals without such a variant.

From a total of 1,006 individuals carrying a NOTCH3-Cys mutations, 102 had an MRI examination, and 96 had all measures available for the present study. These individuals were all heterozygous for a cysteine-altering mutation within the EGFr domain 7-34. To identify NOTCH3-Cys mutation carriers, we extracted bulk exome PLINK files (gaston R package^23^) to search for 99 known cysteine-altering mutations in this gene as already documented by Cho et al.^19^. Among all NOTCH3-Cys carriers, vascular risk factors, prior stroke prevalence and variant distribution across the 34 EGFr domains did not differ by MRI availability (Table S1a, Figure S1).

### Neuroimaging Methods

Brain MRI scans were acquired at three dedicated imaging centers using identical 3T Siemens scanners.^24^ The standardized protocol included T1-weighted MPRAGE (1×1×1 mm³ resolution, TR=2000ms, TE=2.01ms, TI=880ms, flip angle=8°) and T2-weighted FLAIR sequences (1.05×1.0×1.0 mm³ resolution, TR=5000ms, TE=395ms, TI=1800ms).^25^

For WMH quantification, the Brain Intensity AbNormality Classification Algorithm (BIANCA) was used.^26^ It is a fully automated supervised method trained on manually segmented WMH masks from a subset of UKB participants (n=53) with high accuracy (Dice=0.85). The resulting WMH volumes were normalized by the total intracranial volume (TIV) ^27^. All WMH values hereafter are normalized WMH.

For BPF calculation, the FMRIB’s Automated Segmentation Tool (FAST) was used.^28^ FAST generated probability maps for gray matter, white matter, and CSF from T1-weighted images, with BPF calculated as the sum of gray and white matter volumes divided by TIV.

Lacune counts were visually assessed by an experimented neurologist (H.C.) using ITK-SNAP (version 3.8.0)^29^ on T1-weighted images for all 96 carriers. Lacunes were defined on 3DT1-weighted images as 3-15 mm subcortical CSF-filled cavities distinct from dilated perivascular spaces.^1^ All imaging data underwent the UKB’s quality control pipeline.^25^

### Statistical analysis

Multiple linear regression models were used with BPF as the dependent variable and WMH as the primary independent variable of interest to evaluate: (1) the main effect of WMH on BPF, and (2) potential interaction effects between WMH and either age or NOTCH3-Cys mutation status.

Five vascular risk factors were considered as potential covariates when needed: hypercholesterolemia, hypertension, diabetes mellitus, daily alcohol consumption, and smoking history (current or former). They were constructed through integration of multiple data sources within the UKB,^20^ and were assessed at both baseline and at time of MRI acquisition to account for temporal changes in risk factor profiles.^30^

To assess the WMH-BPF relationships, we first implemented population-specific linear regression models (M1) separately for carriers (n=96) and for non-carriers (n=44,847) using sex and age as supplementary covariates. Thereafter, we examined age-dependent effects by fitting M1 in six age strata (≤53 years, ≥74 years, and 4 strata of 5 years each in between) for each group. In carriers, a second model (M2) was derived from M1 including lacune count as an additional covariate.

Finally, to compare WMH-BPF associations between the two groups while accounting for their potential non-linear relationships, we developed a third model (M3) a Generalized Additive Model (GAM) fitted to the combined population incorporating spline terms for age and WMH, binary sex and genetic mutation status, and all significant interactions between age, WMH, sex, and mutation (identified from a preliminary model with all possible two-way interactions). Full specifications of these models are detailed in Table S2. To prevent distortion from the natural fact that carriers may have more WMH than non-carriers, we used a non-linear (spline) WMH term in our model allowing us to properly measure the true difference in WMH effects between groups.

Sensitivity analyses were performed and included: (1) all models with additional adjustment for the vascular risk factors (M4), and (2) comparison of vascular risk factors and stroke outcomes between participants with and those without MRI to assess the sample’s representativeness of the entire corresponding UKB population.

## Results

### Differences between carriers and non-carriers of NOTCH3-Cys variants

Carriers and non-carriers of NOTCH3-Cys variants had similar distributions of sex, age, vascular risk factors, and BPF (Table 1). NOTCH3-Cys carriers were most likely to have a history of prior stroke (p=0.17 for MRI only individuals, Table 1; p=2.10^-4^ for all participants, Table S1-b). NOTCH3-Cys carriers exhibited (p=0.03) a marginally higher mean BPF (81.1%) than non-carriers (80.5%).

**Table 1.** Main demographic, vascular risk factors at imaging date, and imaging results according to the presence or absence of a NOTCH-Cys variant in the UKB individuals with MRI examination. The p-values test the difference between carriers and non-carriers in each variable’s distribution. All the tests are adjusted on age and sex except when the variable being compared is age or sex. Statistically significant results in bold.

|  | NOTCH3-Cys +<br>n = 96 | NOTCH3-Cys -<br>n = 44847 | P-value |
| --- | --- | --- | --- |
| **Demographics** |  |  |  |
| Age, mean ± st. dev. [N] | 64.1 ± 7.3 [96] | 64.3 ± 7.7 [44847] | 0.83 |
| Male sex, n (%) [N] | 47 (49 %) [96] | 21105 (47.1 %) [44847] | 0.71 |
| **Vascular risk factors** |  |  |  |
| Hypertension, n (%) [N] | 62 (72.1 %) [86] | 26297 (64.3 %) [40906] | 0.11 |
| Hypercholesterolemia, n (%) [N] | 44 (48.4 %) [91] | 21823 (51.1 %) [42672] | 0.63 |
| Diabetes, n (%) [N] | 9 (11.5 %) [78] | 2928 (8 %) [36762] | 0.24 |
| History of smoking, n (%) [N] | 47 (49 %) [96] | 23563 (53.1 %) [44372] | 0.4 |
| Daily alcohol use, n (%) [N] | 15 (15.6 %) [96] | 7481 (16.8 %) [44512] | 0.74 |
| **Outcomes** |  |  |  |
| History of stroke, n (%) [N] | 3 (3.1 %) [96] | 649 (1.4 %) [44845] | 0.17 |
| Fluid intelligence score, mean ± st. dev. [N] | 6.9 ± 1.9 [83] | 6.6 ± 2.1 [41211] | 0.19 |
| **Imaging measures** |  |  |  |
| BPF (% of TiV), mean ± st. dev. [N] | **81.1 ± 2.5 [96]** | **80.5 ± 2.9 [44847]** | **0.032** |
| WMH volume (% of TiV), mean ± st. dev. [N] | **0.83 ± 1 [96]** | **0.36 ± 0.5 [44847]** | **p < 10<sup>-8</sup>** |
| Median (range; Q1 - Q3) | 0.47 (0.02-6.70 ; 0.18-1.04) | 0.47 (4.10<sup>-4</sup>-8.03 ; 0.11-0.42) |  |
| Number of lacunes, mean ± st. dev. [N] | 0.5 ± 1.2 [96] | NA | |
| Median (range; Q1 - Q3) | 0 (0 - 6 ; 0 - 0) | NA |  |
| At least one lacune, n (%) [N] | 17 (17.7 %) [96] | NA |  |

In contrast, the two groups demonstrated (p < 10^-8^) markedly different distributions of WMH, with carriers showing a mean WMH volume more than twice that of non-carriers (0.83% vs 0.36%). Due to the larger sample size of non-carriers, the range of WMH volumes in carriers was nevertheless entirely encompassed within that of non-carriers, with a subset of individuals exhibiting values in the high single-digit percentages (Figure 1). In both carriers and non-carriers, WMH volume correlated positively with age, whereas BPF showed a strong negative correlation with age (Table S3).

**Figure 1.**
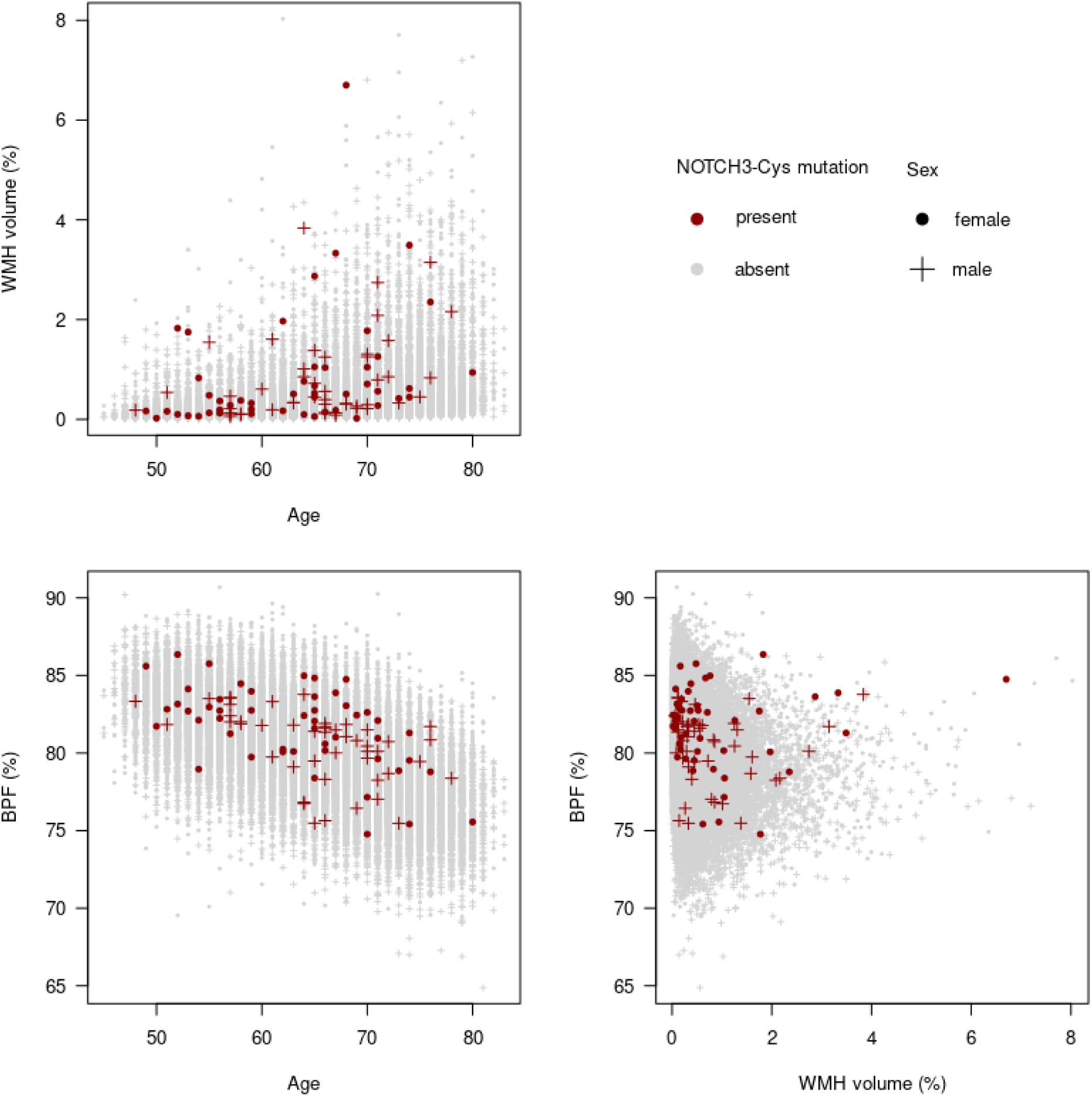
Distribution of WMH and BPF measured on MRI according to age in the UK Biobank. Individuals with a NOTCH3-Cys variant were isolated (red) from the rest of the population (grey). See table S3 for the associated correlation coefficients.

### Relationships between WMH and BPF in non-carriers of NOTCH3-Cys variants

In non-carriers, the age- and sex-adjusted linear regression model (M1-nc) showed a significant inverse association between WMH volume and BPF (Table 2A). For each 1% increase in WMH volume, BPF decreased by 0.116% on average (β=-0.116, p=3 × 10⁻⁶; n=44,847) at constant age and sex. This association was highly statistically significant (p=3*10^-6^) but the corresponding effect size was limited (in regards to the large sample size i.e., n=44,847): adding WMH to the model increased the R² from 0.3719 to 0.3722 compared with a model including only age and sex.

**Table 2.**
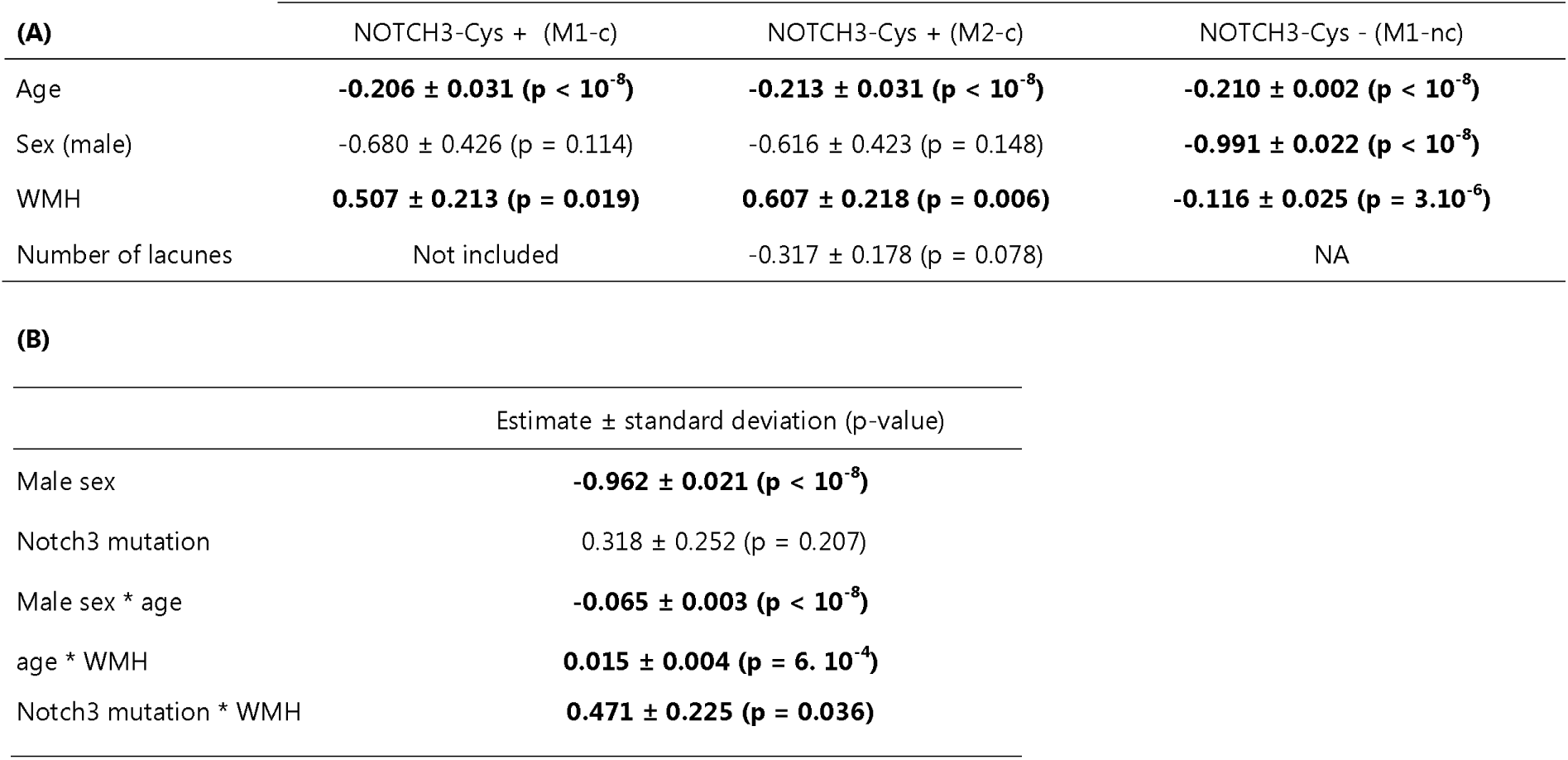
Regression Coefficient Estimates (A) of the final linear models M1-c, M2-c, M1-nc used for separate analysis of UK Biobank individuals having MRI data, and (B) of the linear terms in the GAM model M3. Coefficient estimates ± standard deviations (p-values) of the three linear models M1-c, M2-c, M1-nc and the linear terms of GAM model M3, with statistically significant results in bold.

| (A) | NOTCH3-Cys + (M1-c) | NOTCH3-Cys + (M2-c) | NOTCH3-Cys - (M1-nc) |
| --- | --- | --- | --- |
| Age | **-0.206 ± 0.031 (p < 10<sup>-8</sup>)** | **-0.213 ± 0.031 (p < 10<sup>-8</sup>)** | **-0.210 ± 0.002 (p < 10<sup>-8</sup>)** |
| Sex (male) | -0.680 ± 0.426 (p = 0.114) | -0.616 ± 0.423 (p = 0.148) | **-0.991 ± 0.022 (p < 10<sup>-8</sup>)** |
| WMH | **0.507 ± 0.213 (p = 0.019)** | **0.607 ± 0.218 (p = 0.006)** | **-0.116 ± 0.025 (p = 3.10<sup>-6</sup>)** |
| Number of lacunes | Not included | -0.317 ± 0.178 (p = 0.078) | NA |

(B)
| | Estimate ± standard deviation (p-value) |
| --- | --- |
| Male sex | **-0.962 ± 0.021 (p < 10<sup>-8</sup>)** |
| Notch3 mutation | 0.318 ± 0.252 (p = 0.207) |
| Male sex * age | **-0.065 ± 0.003 (p < 10<sup>-8</sup>)** |
| age * WMH | **0.015 ± 0.004 (p = 6. 10<sup>-4</sup>)** |
| Notch3 mutation * WMH | **0.471 ± 0.225 (p = 0.036)** |

In contrast, BPF exhibited a strong negative association with age, with BPF declining by 0.21% per year when WMH was held constant. Women also had a higher mean BPF than men by 0.99% on average. Adjustment for vascular risk factors had minimal impact on the association between WMH and BPF (Table S4).

To further explore age-specific effects, we stratified the non-carrier sample into six 5-year age bins (except the youngest and oldest bins) and refit the linear model (M1-nc) within each stratum (n=4,600-10,000 per bin). The resulting β-WMH estimates, corresponding to the WMH effects on BPF at constant age, differed according to age strata in support for a variable age-dependent pattern of aging (Figure 2). At younger age, WMH was associated with brain atrophy (β-WMH ≈ -0.5% decrease of BPF per 1% WMH increase), but this effect decreased progressively and vanished, reaching near-zero at the oldest age range.

**Figure 2.**
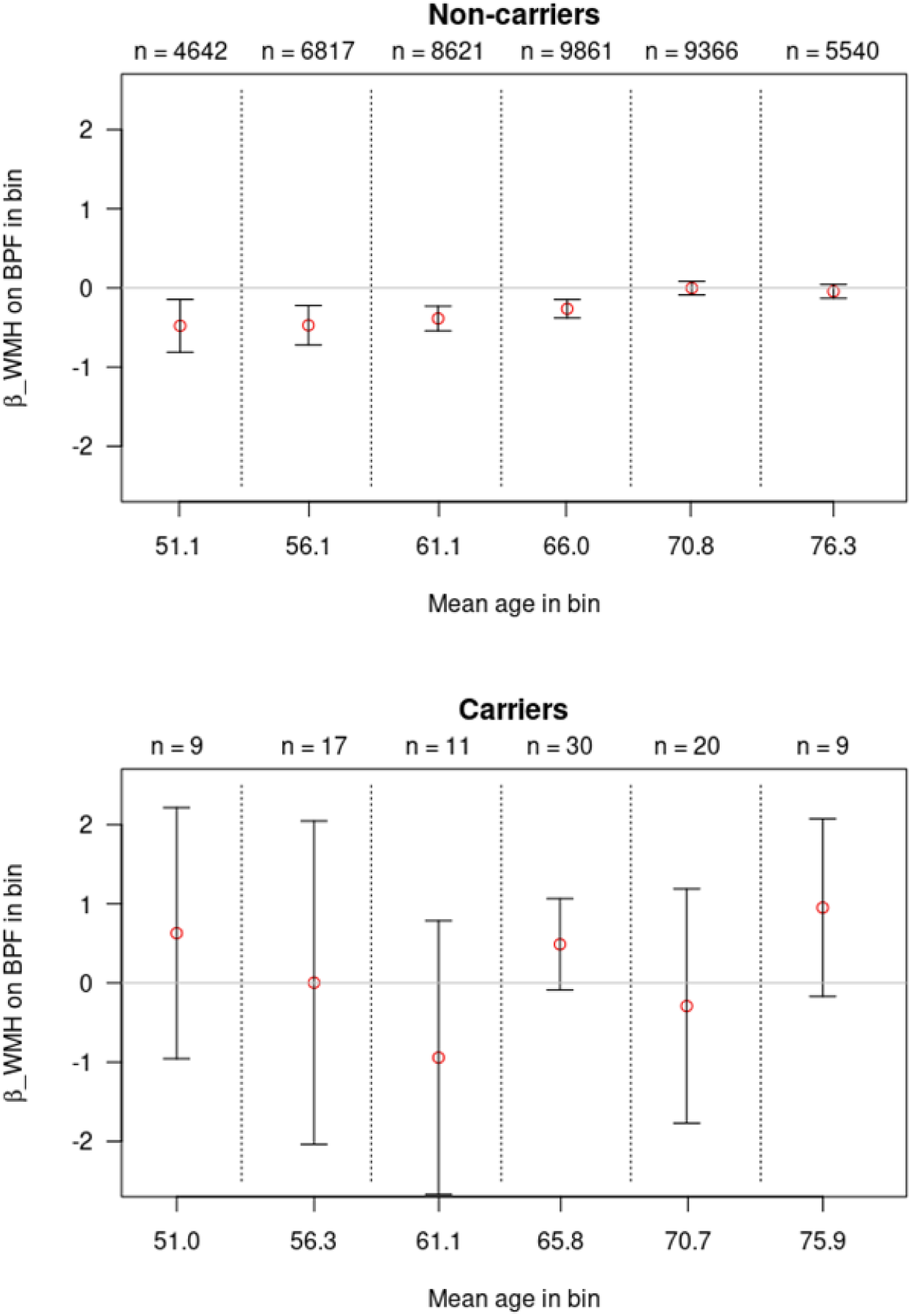
Age-Stratified Association Between White Matter Hyperintensity Burden and Brain Parenchymal Fraction. The figure presents estimates of the linear effect of white matter hyperintensity (WMH) volume on brain parenchymal fraction (BPF) (β_WMH_), expressed as percentage change in BPF per percentage change in WMH, across six age bins in NOTCH3-Cys carriers and non-carriers. Dots represent point estimates of β_WMH_, vertical bars indicate 95% confidence intervals, and dotted vertical lines mark age bin boundaries. Sample sizes for each bin appear at the top of the panels. The two panels use different y-axis scales. The effect of WMH on BPF shifts from significantly negative at younger ages to null at older ages. β_WMH_ remains stable below age 60 at approximately -0.5% BPF decrease per 1% WMH increase, it increases markedly between ages 60 and 70 years and stabilizes around 0 for ages >70 years. In carriers, the sample sizes by bin are too low to get precise estimates of the β_WMH_.

### Relationships between WMH and BPF in carriers of NOTCH3-Cys variants

Linear regression (Model M1-c, Table 2A) showed that, unlike in non-carriers, NOTCH3-Cys carriers exhibited a positive association between WMH and BPF after adjustment for age and sex (β=0.51% increase of BPF per 1% WMH volume; p=0.019). Including WMH in the model improved the fit (R²=0.3645 vs. 0.3254 for age and sex alone).

Age remained strongly and negatively associated with BPF (β=−0.21% per year; p<0.001), while the sex-BPF relationship did not remain significant. Adjustment for vascular risk factors in the same model did not meaningfully alter the positive association between WMH and BPF (Table S4).

Adjustment for lacunes numbers (Model M2-c, Table 2A) increased the association strength between WMH and BPF (β=0.61% increase of BPF per 1% increase of WMH at constant age; p=0.006). Lacunes counts was positively associated with WMH and negatively associated with BPF, suggesting partial mediation. However, among carriers of the NOTCH3-Cys variants, the direct positive association between WMH and BPF remained dominant.

To assess age dependency of the WMH-BPF association in carriers of NOTCH3-Cys variants, the sample was also divided into six age bins as previously done in non-carriers (Figure 2). The sample size of the resulting bins was small (bins of 6-30 individuals). Based on these data, no significant age-related change in the relationship between WMH and BPF was detected (Figure 2).

### Comparison of WMH-BPF association by NOTCH3-Cys Variant Status

To assess the modifying effect of the NOTCH3-Cys variant on the WMH-BPF relationship, we applied a GAM (Model M3, Table S2) to the full MRI data cohort. The model estimated BPF as a function of non-linear spline terms for age and WMH, binary effects of sex and NOTCH3-Cys carrier status, and three significant interaction terms identified in preliminary analyses: NOTCH3-Cys × WMH, age × WMH, and age × sex (Table S2).

The NOTCH3-Cys × WMH interaction (P=0.036) confirmed that the association between WMH and BPF differed significantly between carriers and non-carriers, consistent with prior stratified analyses (Table 2B). This difference persisted even when allowing non-linear WMH effects in both groups, with the interaction modelled as a linear deviation from a shared non-linear WMH component. In non-carriers, the WMH-BPF relationship was modestly negative and convex, whereas in NOTCH3-Cys carriers, the same non-linear function was augmented by a steeply positive linear component, yielding near-neutral effects at low WMH volumes and increasingly positive effects at higher volumes (Figure 3).

**Figure 3.**
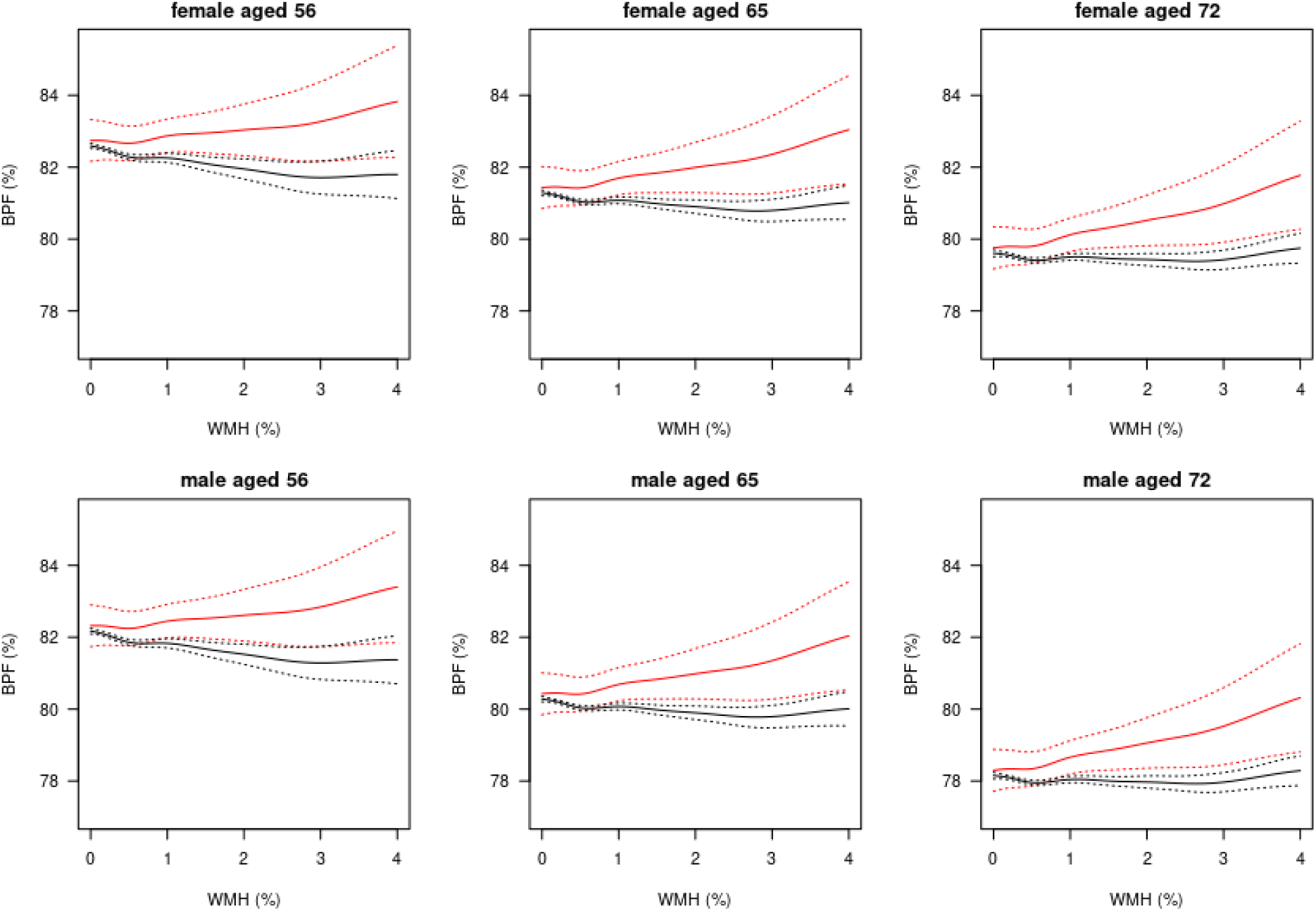
Spline-Based Estimates of Brain Parenchymal Fraction (BPF) as a Function of White Matter Hyperintensity Burden (WMH). Estimated BPF as a spline function of WMH depending on age, sex and NOTCH3-Cys mutation presence (carriers in red). Dotted lines are 95% confidence intervals. The range of WMH from 0% to 4% covers 99% of the sample of carriers and 99.8% of the non-carriers. Three fixed ages are considered: 65 is median age of both carriers and non-carriers, and Q1 = 56 and Q5 = 72 are age quantiles of non-carriers at 1/6 and 5/6.

A highly significant age × sex interaction (p<10⁻⁸) indicated that the age-related BPF decline was more pronounced in men than women, independent of WMH burden (Figure S2). The age × WMH interaction (p=5.7×10⁻⁴) further demonstrated that the WMH-BPF relationship varied across ages, corroborating findings from age-binned linear regressions (Figure 2) without assuming linearity. Adjustment for vascular risk factors attenuated the NOTCH3-Cys × WMH interaction (β=+0.47 to +0.44) and reduced its statistical significance (P=0.036 to 0.076), likely due to reduced power from additional inclusion of covariates.

## Discussion

This population-based neuroimaging investigation demonstrates markedly different relationships between WMH and BPF in individuals harboring NOTCH3-Cys variants compared with non-carriers, providing new insights into the natural history of cSVD.^31^ Unlike non-carriers, carriers showed a robust positive association between WMH volume and BPF, consistent across age groups and strengthened after adjustment for lacune count. This contrasts with the negative WMH-BPF relationship observed in individuals without the variant^1,32^ and supports that NOTCH3-related SVD represents a distinct form of microvascular pathology within the spectrum of SVD detected in the general population.^9,10^ These findings also further support that SVD is not a uniform entity at population level, but comprises biologically distinct subtypes with different structural trajectories, already detectable at early or subclinical stages.^33^

NOTCH3-Cys variants carriers displayed larger WMH volumes than non-carriers after adjustment for age and vascular risk factors, but their WMH burden remained substantially lower than that reported in clinical CADASIL cohorts.^10,34^ Median WMH volume is approximately fivefold lower than in symptomatic individuals.^35^ These results indicate that this imaging subgroup largely represents preclinical or mild disease stages. Importantly, the 100-fold difference between the population frequency of NOTCH3-Cys variants of ∼1 in 467 already reported in the UKB^31^ and the much lower prevalence of clinically diagnosed CADASIL patients in the literature (2-5 per 100,000)^10,34^ reinforces the notion that most carriers in the UKB can remain asymptomatic or develop only minimal white matter changes. Thus, population screening captures a much earlier disease spectrum than clinical cohorts, offering a unique opportunity to study structural correlates of such genetically determined SVD from onset of WMH.^9,31^

The divergent WMH-BPF relationships observed between carriers and non-carriers underscore that white matter changes do not reflect a uniform pathological process in SVD.^1^ In non-carriers, the modest negative association between WMH and BPF was strongest among younger participants and weakened with age, likely because global atrophy increases in older adults and masks the effects of WMH.^32^ This age-dependent effect is corroborated by the Rotterdam Scan Study, where the WMH-brain volume association weakened significantly with age, supporting the masking effect of global atrophy.^7,32^ In contrast, the positive association in NOTCH3-Cys variant carriers remained consistent across age groups and became stronger when accounting for lacune count, suggesting that WMH and lacunes may represent distinct pathological pathways during disease progression.^10,36^ The robustness of this association across separate analyses, and the significant genetic interaction observed in additive modelling, reinforce the conclusion that NOTCH3 variants modify the fundamental relationship between lesion accumulation and brain volume.^31^ Despite the relatively small WMH burden in the UKB, this positive relationship in variant carriers aligns with previous reports that WMH may reflect heterogeneous tissue states in CADASIL, some representing tissue water accumulation rather than demyelination or axonal loss.^37^ The observed findings are consistent with emerging evidence that NOTCH3-related pathology could involve astrocytic dysfunction, dysregulation of water homeostasis, and dilation of perivascular spaces, driven by accumulation of the NOTCH3 extracellular domain (ECD) in small cerebral vessels.^38,39^. Whether alterations of aquaporin channels on astrocytic end-feet already reported post-mortem are involved since the pre-symptomatic stage or at the emergence of WMH in presence of different NOTCH-Cys variants remains to be determined.^40^ The positive WMH-BPF association likely represents a transient phase in which lesions correspond to increased water content before evolving into the tissue loss seen and severe atrophy detected in most advanced CADASIL cases.^10^ Whether these effects result from lower levels of NOTCH3-ECD accumulation, variant-specific molecular properties, or other genetic/environmental modifiers remains unclear.^38,41^

Several limitations must be considered. The cross-sectional design precludes causal or temporal inferences, and it remains unknown whether the positive WMH-BPF relationship can transition to the negative association with aging.^10^ The relatively small number of NOTCH3-Cys variant carriers limits statistical power to detect subtle modifiers, though the consistency of findings across models provides reassurance. While adjustments were made for major vascular risk factors, residual confounding from unmeasured variables cannot be excluded. Furthermore, the UKB’s healthy volunteer bias^21^ and imaging selection biases^25^ limit generalizability to more severely affected populations. Nevertheless, the study has notable strengths, including its large sample size, detailed phenotyping, and standardized neuroimaging protocols.^24^ The use of advanced statistical modelling allowed identification of non-linear and genotype-specific effects that conventional approaches might miss.^31^ Importantly, the population-based design provides access to the preclinical stage of NOTCH3-Cys variant related biology that remains largely invisible in clinical cohorts, offering insight into the earliest manifestations of NOTCH3-ECD accumulation.^9^

Overall, this study illustrates that WMH–brain volume relationships are biologically context-dependent in SVD and must be interpreted within a genetic and disease-stage framework, as their direction may evolve differently with aging and lesion accumulation depending on genetic background.

## Data Availability

The individual-level data used in this study were obtained from UK Biobank under Application No. 94113. UK Biobank data are available to eligible bona fide researchers for health-related research in the public interest through the UK Biobank data access procedures. These data are not publicly available without restriction.

## Data availability

The data are from UK Biosbank. UK Biobank is only accessible through individual researcher application and cannot be shared directly.

## Acknowledgements

This research has been conducted using the UK Biobank Resource under Application Number 94113. We thank Thérèse Truong, Eva Jaber and Frédéric Robergeau for their help with the UK Biobank data and Louis Lambert for general discussions.

