## Supplemental file for "NOTCH3 cysteine variants alter the white matter lesion-brain volume link in the general population"

### Table S1-a

**Title:** Comparison of Baseline Vascular Risk Factors Between Participants With and Without Brain MRI

**Legend:** Baseline vascular risk factors according to subsequent brain MRI availability (with WMH and BPF data). Except for age and fluid intelligence score, values are expressed as number of cases (percentage) [sample size]. The p-values test whether a variable's distribution differs at baseline according to subsequent MRI availability, within each mutation status. All the tests are adjusted on age and sex except when the variable being compared is age or sex.

| | NOTCH3-Cys + | | | NOTCH3-Cys - | | |
|---|---|---|---|---|---|---|
| | with brain MRI n = 96 | without brain MRI n = 910 | p-value | with brain MRI n = 44847 | without brain MRI n = 456086 | p-value |
| **Demographics** | | | | | | |
| Age, mean ± st. dev. [n] | 55 ± 7 [96] | 56.1 ± 8 [910] | 0.21 | 55 ± 7.5 [44847] | 56.7 ± 8.1 [456086] | $< 10^{-10}$ |
| Male sex | 47 (49 %) [96] | 443 (48.7 %) [910] | 0.96 | 21105 (47.1 %) [44847] | 207305 (45.5 %) [456086] | $< 10^{-10}$ |
| **Risk factors** | | | | | | |
| Hypertension | 51 (53.1 %) [96] | 508 (55.8 %) [910] | 0.87 | 22690 (50.6 %) [44831] | 274972 (60.4 %) [455095] | $< 10^{-10}$ |
| Hypercholesterolemia | 42 (45.7 %) [92] | 466 (52.7 %) [884] | 0.34 | 18956 (44.9 %) [42255] | 220944 (51.1 %) [432078] | $< 10^{-10}$ |
| Diabetes | 5 (6.4 %) [78] | 85 (10.9 %) [781] | 0.27 | 2154 (5.9 %) [36597] | 40409 (10.7 %) [376456] | $< 10^{-10}$ |
| Tobacco | 50 (52.1 %) [96] | 513 (56.7 %) [905] | 0.39 | 25497 (57 %) [44748] | 272354 (60.1 %) [453306] | $< 10^{-10}$ |
| Daily alcohol | 18 (18.8 %) [96] | 168 (18.5 %) [906] | 0.92 | 10111 (22.6 %) [44824] | 91379 (20.1 %) [454613] | $< 10^{-10}$ |
| **Outcomes** | | | | | | |
| Stroke | 2 (2.1 %) [96] | 29 (3.2 %) [910] | 0.68 | 330 (0.7 %) [44847] | 7761 (1.7 %) [456086] | $< 10^{-10}$ |
| Fluid intelligence score, mean ± st. dev. [n] | 6.2 ± 1.9 [29] | 5.8 ± 2.3 [291] | 0.29 | 6.7 ± 2 [15083] | 5.9 ± 2.2 [149883] | $< 10^{-10}$ |

**Table S1-b**

**Title:** Comparison of Baseline Vascular Risk Factors Between Notch3-Cys carriers and non-carriers

**Legend:** Baseline vascular risk factors according to Notch3-Cys status, regardless of subsequent brain MRI availability. The p-values test whether a variable's distribution differs at baseline between carriers and non-carriers. All the tests are adjusted on age and sex except when the variable being compared is age or sex.

| Data | NOTCH3-Cys + n = 1006 | NOTCH3-Cys - n = 500933 | p-value |
|---|---|---|---|
| **Demographics** | | | |
| Age, mean ± st. dev. [N] | 56 ± 7.9 [1006] | 56.5 ± 8.1 [500933] | 0.022 |
| Male sex, n (%) [N] | 490 (48.7 %) [1006] | 228410 (45.6 %) [500933] | 0.048 |
| **Vascular risk factors** | | | |
| Hypertension, n (%) [N] | 559 (55.6 %) [1006] | 297662 (59.5 %) [499926] | 0.023 |
| Hypercholesterolemia, n (%) [N] | 508 (52 %) [976] | 239900 (50.6 %) [474333] | 0.096 |
| Diabetes, n (%) [N] | 90 (10.5 %) [859] | 42563 (10.3 %) [413053] | 0.76 |
| History of smoking, n (%) [N] | 563 (56.2 %) [1001] | 297851 (59.8 %) [498054] | 0.017 |
| Daily alcohol use, n (%) [N] | 186 (18.6 %) [1002] | 101490 (20.3 %) [499437] | 0.15 |
| **Outcomes** | | | |
| History of stroke, n (%) [N] | 31 (3.1 %) [1006] | 8091 (1.6 %) [500933] | $2.10^{-4}$ |
| Fluid intelligence score, mean ± st. dev. [N] | 5.8 ± 2.3 [320] | 6 ± 2.2 [164966] | 0.12 |

**Table S2**

**Title:** Mathematical expression of the different linear models used in carriers of Notch3Cys variant (group c) and non-carriers (nc) of this variant

| |
|---|
| **Models M1-c and M1-nc**<br>with no interaction terms performed separately in c and nc<br>$BPF = \beta_{sex} 1_{male} + \beta_{age} age + \beta_{WMH} WMH + \epsilon$ |
| **Model M2-c**<br>with additional adjustment for the number of lacunes performed in group c<br>$BPF = \beta_{sex} 1_{male} + \beta_{age} age + \beta_{WMH} WMH + \beta_{Lacunes} N_{Lacunes} + \epsilon$ |
| **Model M3**<br>with splines and interactions in both c and nc<br>$BPF = \beta_{sex} 1_{male} + S_{age}(age) + S_{WMH}(WMH) + \beta_{notch3} 1_{notch3}$<br>$+ \beta_{sex*age} 1_{male} age + \beta_{age*WMH}\ age * WMH + \beta_{notch3*WMH} 1_{notch3} WMH + \epsilon$ |
| **Model M4**<br>with splines and interactions in c and nc and significant interaction in M3<br>$BPF = \beta_{sex} 1_{male} + S_{age}(age) + S_{WMH}(WMH) + \beta_{notch3} 1_{notch3}$<br>$+ \beta_{sex*age} 1_{male} age + \beta_{sex*WMH} 1_{male} WMH + \beta_{sex*notch3} 1_{male} 1_{notch3}$<br>$+\beta_{age*WMH}\ age * WMH + \beta_{notch3*age}\ 1_{notch3} age + \beta_{notch3*WMH} 1_{notch3} WMH + \epsilon$ |

**Table S3**

**Title**: Linear correlation coefficients of WMH and BPF with age among Notch3-Cys carriers and non-carriers.

**Legend**: The correlation coefficient estimates are displayed with their 95% confidence intervals. These correlations are visible in the scatterplots of Figure 1.

| | **NOTCH3-Cys +**<br>n = 96 | **NOTCH3-Cys -**<br>n = 44943 |
|---|---|---|
| ρ(Age, WMH) | 0.31 [0.11, 0.48] | 0.383 [0.375, 0.391] |
| ρ(Age, BPF) | - 0.550 [-0.68,-0.39] | - 0.585 [-0.591, -0.579] |

**Table S4**

**Title**: Regression results obtained in samples of individuals with the NOTCH3-Cys variant with and without adjustment for lacunes and in individuals without the variant in the UK Biobank

**Legend**: Coefficient estimates ± standard deviations (p-values) of the three linear models, with statistically significant results in bold.

| Model / Variable | Noth3-cys + | Noth3-cys + (with adjustment by lacunes) | Noth3-cys - |
|---|---|---|---|
| Age | **-0.188 ± 0.036 ($p = 3.10^{-6}$)** | **-0.191 ± 0.037 ($p = 3.10^{-6}$)** | **-0.206 ± 0.002 ($p < 10^{-8}$)** |
| Sex (male) | -0.261 ± 0.542 (p = 0.632) | -0.232 ± 0.546 (p = 0.673) | **-0.961 ± 0.025 ($p < 10^{-8}$)** |
| Hypercholesterolemia | **-1.315 ± 0.522 (p = 0.014)** | **-1.316 ± 0.524 (p = 0.015)** | **0.059 ± 0.026 (p = 0.026)** |
| Hypertension | 0.248 ± 0.587 (p = 0.674) | 0.243 ± 0.59 (p = 0.682) | 0.04 ± 0.028 (p = 0.149) |
| Diabetes | -0.462 ± 0.879 (p = 0.601) | -0.338 ± 0.901 (p = 0.709) | **-0.74 ± 0.047 ($p < 10^{-8}$)** |
| Smoking history | 0.277 ± 0.511 (p = 0.59) | 0.247 ± 0.515 (p = 0.633) | **-0.154 ± 0.025 ($p < 10^{-8}$)** |
| Daily use of alcohol | -0.199 ± 0.725 (p = 0.784) | -0.16 ± 0.73 (p = 0.828) | **-0.446 ± 0.034 ($p = 3.10^{-4}$)** |
| WMH | 0.509 ± 0.238 (p = 0.037) | 0.553 ± 0.248 (p = 0.03) | **-0.102 ± 0.028 ($p < 10^{-8}$)** |
| Number of lacunes | Not included | -0.154 ± 0.223 (p = 0.494) | **NA** |

**Figure S1**

**Title:** Number of NOTCH3-Cys mutation carriers by EGFR domain of their mutation, comparing individuals with or without MRI.

**Legend:** The scales' ratio is such that if there is perfect proportionality of carrier numbers in an EGFR domain, the red dot for that EGFR is attached at the top of black bar. R1231C (abbreviated RC) and C1222G (abbreviated CG) are counted separately from the other EGFR 31 mutations and therefore excluded from the column 31. A Chi-square test found no significant deviation from proportionality between the two samples (p = 0.62).

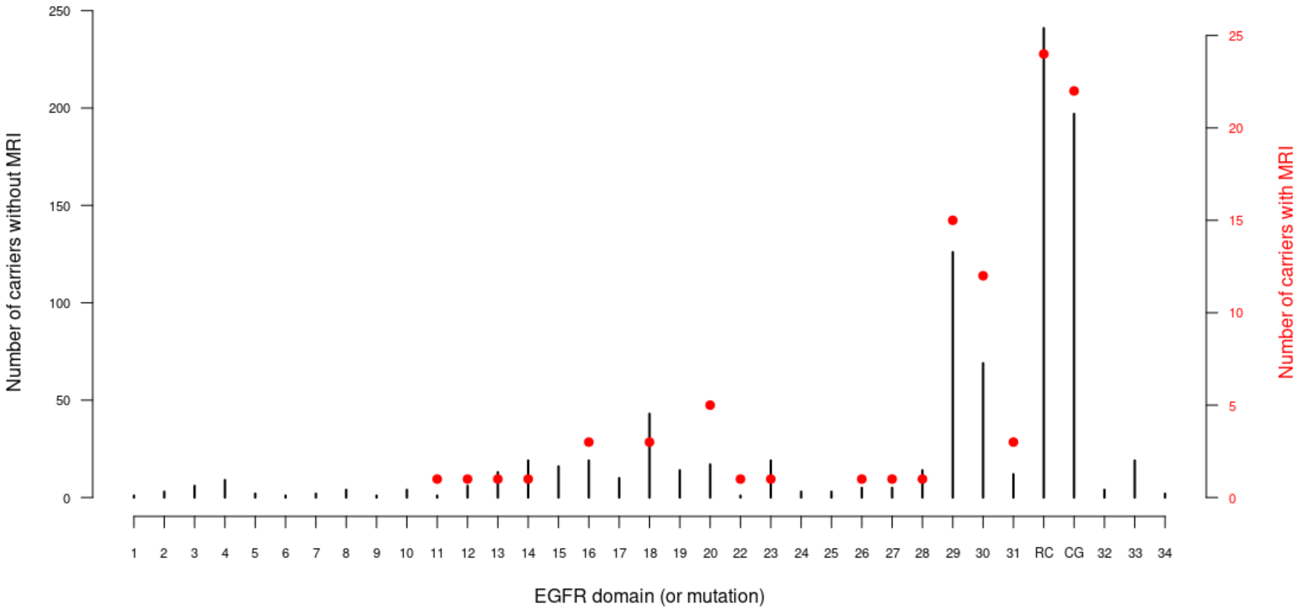

**Figure S2.**

**Title**: Estimated brain parenchymal fraction (BPF) modelled as a spline function of age in male and female non-carriers of the NOTCH3-Cys variant assuming there was no white matter hyperintensities (WMH = 0%).

**Legend**: The curves represent sex-specific spline estimates of BPF across age, illustrating the expected age-related trajectory of brain parenchymal fraction in the absence of WMH burden.

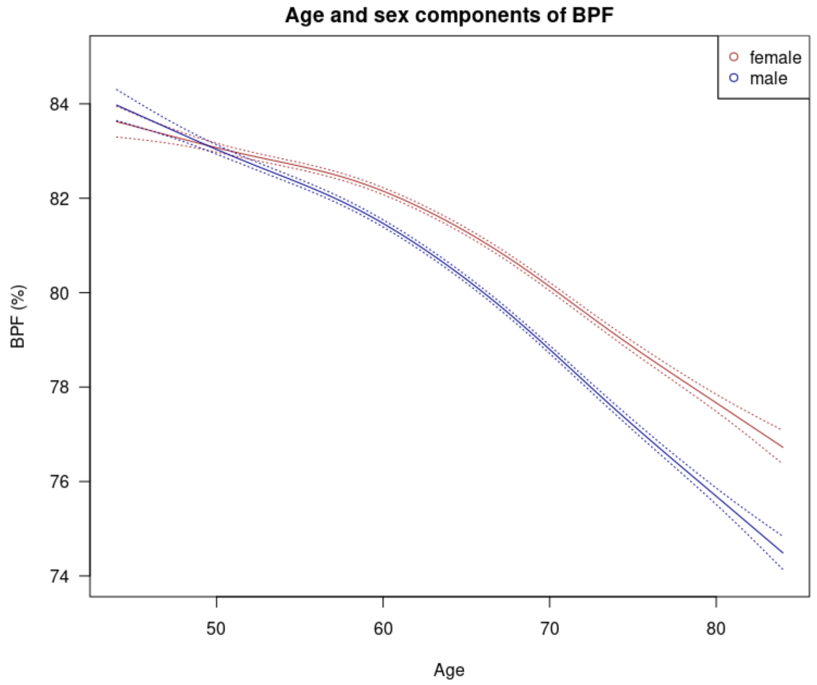
